# Multimodal Digital Phenotyping of Behavior in a Neurology Clinic: Development of the Neurobooth Platform and the First Two Years of Data Collection

**DOI:** 10.1101/2024.12.28.24319527

**Authors:** Adonay S. Nunes, Siddharth Patel, Brandon Oubre, Mainak Jas, Divya D. Kulkarni, Anna C. Luddy, Nicole M. Eklund, Faye X. Yang, Rohin Manohar, Nancy N. Soja, Katherine M. Burke, Bonnie Wong, Dmitry Isaev, Steven Espinosa, Jeremy D. Schmahmann, Christopher D. Stephen, Anne-Marie Wills, Albert Hung, Bradford C. Dickerson, James D. Berry, Steven E. Arnold, Vikram Khurana, Lawrence White, Guillermo Sapiro, Krzysztof Z. Gajos, Sheraz Khan, Anoopum S. Gupta

**Affiliations:** Department of Neurology, Massachusetts General Hospital, Harvard Medical School, Boston, MA, USA; Martinos Center for Biomedical Imaging, Massachusetts General Hospital, Harvard Medical School, Boston, MA, USA; Massachusetts General Hospital Institute of Health Professions, Boston, MA, USA; Department of Psychiatry, Massachusetts General Hospital, Harvard Medical School, Boston, MA, USA; Department of Biomedical Engineering, Duke University, Durham, NC, USA; Department of Electrical and Computer Engineering, Duke University, Durham, NC, USA; Department of Neurology, Brigham and Women’s Hospital, Harvard Medical School, Boston, MA, USA; Department of Electrical and Computer Engineering, Princeton University, NJ, USA; Harvard John A. Paulson School of Engineering and Applied Sciences, Allston, Massachusetts, USA

## Abstract

Quantitative analysis of human behavior is critical for objective characterization of neurological phenotypes, early detection of neurodegenerative diseases, and development of more sensitive measures of disease progression to support clinical trials and translation of new therapies into clinical practice. Sophisticated computational modeling can support these objectives, but requires large, information-rich data sets. This work introduces Neurobooth, a customizable platform for time-synchronized multimodal capture of human behavior. Over a two year period, a Neurobooth implementation integrated into a clinical setting facilitated data collection across multiple behavioral domains from a cohort of 470 individuals (82 controls and 388 with neurologic diseases) who participated in a collective 782 sessions. Visualization of the multimodal time series data demonstrates the presence of rich phenotypic signs across a range of diseases. These data and the open-source platform offer potential for advancing our understanding of neurological diseases and facilitating therapy development, and may be a valuable resource for related fields that study human behavior.

## Main Text

Neurological disorders are the leading cause of disability and the second leading cause of death worldwide^1^. There is a large unmet need in neurology for safe and effective therapies. Drug development efforts for neurodegenerative diseases have accelerated in recent years^2^. However, efficient evaluation of new therapies is limited by subjective and insensitive outcome measures, diagnostic uncertainty, incomplete patient stratification, and difficulty in evaluating therapies at the earliest stages of disease^3–5^. Quantitative characterization of digitized behavior using signal processing and machine learning approaches has the potential to address these limitations and accelerate neurology drug development^6^.

Through their effects at the molecular, cellular, and circuit levels, neurodegenerative diseases progressively alter motor and cognitive behavior in life-changing and characteristic ways. These behavioral changes are carefully evaluated by the neurologist during a neurological examination^7^ in order to 1) identify affected components of the nervous system, 2) make an accurate diagnosis, 3) track disease progression, and 4) evaluate response to interventions. Although powerful, clinician-performed assessments cannot capture behavioral patterns that exist beyond the threshold of human perception or that manifest during naturalistic behaviors not represented in the neurological examination. There is also information loss between what the clinician observes and the data recorded, which limits the ability to produce granular datasets to support discovery of new behavioral patterns and identification of disease subgroups. Quantitative capture of behavioral data has the potential to generate information-rich datasets and enable the use of sophisticated, data-driven computational methods to identify clinically-relevant behavioral patterns^6,8^. Adoption of standardized platforms for digital capture of behavior could lead to large scale datasets for training advanced, data-driven models that could broadly transform our understanding of neurological disease and human behavior.

There is a long history of quantitative behavioral data collection in neurological diseases. Various sensor systems have been used to record and assess eye movements using video oculography^9–16^, speech using microphones^17–21^, and body movement using wearable sensors^22–28^ and motion capture^29–32^ systems. It has been shown that eye trackers can detect eye movement control alterations in neurodegenerative populations at early stages^11,33–35^. Computer vision-based analysis of video data obtained from consumer-grade cameras can be used to characterize and quantify gait, hand movements, and eye movements from individuals with neurological diseases^36–41^. Wearable devices have been widely used in neurological populations to quantify gait and limb movements, and to perform disease classification and estimate severity^42–46^. Computational analysis of speech has demonstrated the ability to detect early disease signs, estimate disease severity, and sensitively measure change in neurodegenerative diseases^47–56^. Finally, it has been shown that quantitative analysis of computer mouse movements in both free-living conditions^57^ and during motor tasks^58–60^ reveals patterns informative for detecting neurodegenerative diseases, including Alzheimer’s, Parkinson’s, and cerebellar ataxias. These studies have improved our understanding of how neurological conditions affect behavior. Despite the value these sensor-based systems have demonstrated in the collection and analysis of quantitative behavioral data, digital measures have not been widely adopted in clinical care or in clinical trials^23,61,62^.

Reasons for the limited impact of digital measures to date are that quantitative phenotyping studies typically involve a small number of participants with a well-established diagnosis, focus on a single behavioral domain, and/or use specialized systems for data collection and analysis. There is concern that measures developed from data collected using specialized systems (i.e., custom devices and task protocols) at a single site in a small number of individuals, will not generalize to the larger population and may be too time consuming and/or complex to implement in a clinical setting or in a multi-site clinical trial. The single population, single domain approach also limits the scientific questions that can be advanced and the computational methods that can be used to discover useful disease patterns in typically heterogeneous neurological populations. For example, to adequately detect and measure behavioral patterns that reflect subtle parkinsonism and spasticity in spinocerebellar ataxia populations, conditions with overt parkinsonism and spasticity, such as idiopathic Parkinson’s disease and hereditary spastic paraplegias, should be included and jointly analyzed.

To address these gaps, we developed a high-throughput, open source, and flexible platform called “Neurobooth” to support large-scale multimodal data collection. We created and deployed the first version of this system in a space embedded within the Massachusetts General Hospital outpatient neurology clinic with integration into the clinical workflow. Over a two-year period, synchronized video, audio, inertial sensor, eye tracker, computer mouse, and button-board data were collected in a standardized manner across 782 sessions from 470 unique participants with wide ranging neurological diagnoses. Here we provide an overview of the software and hardware platform and the data generated along with data visualizations. This platform is available to researchers to support quantitative behavioral phenotyping studies and the dataset can be requested. Finally, this study demonstrates the feasibility to integrate into a clinical workflow, providing an avenue for much larger scale data generation with future implementation at additional clinical locations. Integration within a clinic also makes it possible to share results with clinicians and patients at the time of the assessment to provide new information and gain feedback.

## Results

### System Capabilities

Neurobooth is an integrated software and hardware system for collecting quantitative behavioral data. It supports presentation of behavioral stimuli and synchronized multimodal data collection across one or more computers. Because the Neurobooth operating system (NeuroboothOS) can be distributed across multiple computers, it enables concurrent, high-throughput recording across a large number of connected devices. Neurobooth software is open source and highly customizable. It is designed to facilitate the addition of new sensors, stimuli, and study parameters, thereby supporting multiple experimental protocols and disease populations. The Neurobooth software also supports the organization, visualization, and analysis of behavioral time series data. These capabilities aim to balance flexibility (to improve and adapt the system) with standardization (to promote data aggregation and replication of results).

### Dataset

The first Neurobooth system (Fig. 1) was deployed in the Massachusetts General Hospital (MGH) neurology clinic in April 2022, supported by an award from the Massachusetts Life Sciences Center. All participants provided informed consent and the research protocol was approved by the MGH Institutional Review Board (2021P000257, approved March 3, 2021). The deployment at MGH aimed to collect longitudinal digital data from a broad range of neurodegenerative disease populations, including presymptomatic individuals who carry a specific disease gene but do not yet have symptoms or definite clinical signs of disease, and individuals who have not yet received a conclusive clinical diagnosis. With MGH Neurobooth situated in the clinic, most participants with neurologic disorders participated in the study directly before or after their regularly scheduled clinic visits, enabling scalable longitudinal data collection. The MGH Neurobooth is also wheelchair accessible to support data collection in disease populations with limited mobility.

**Figure 1.**
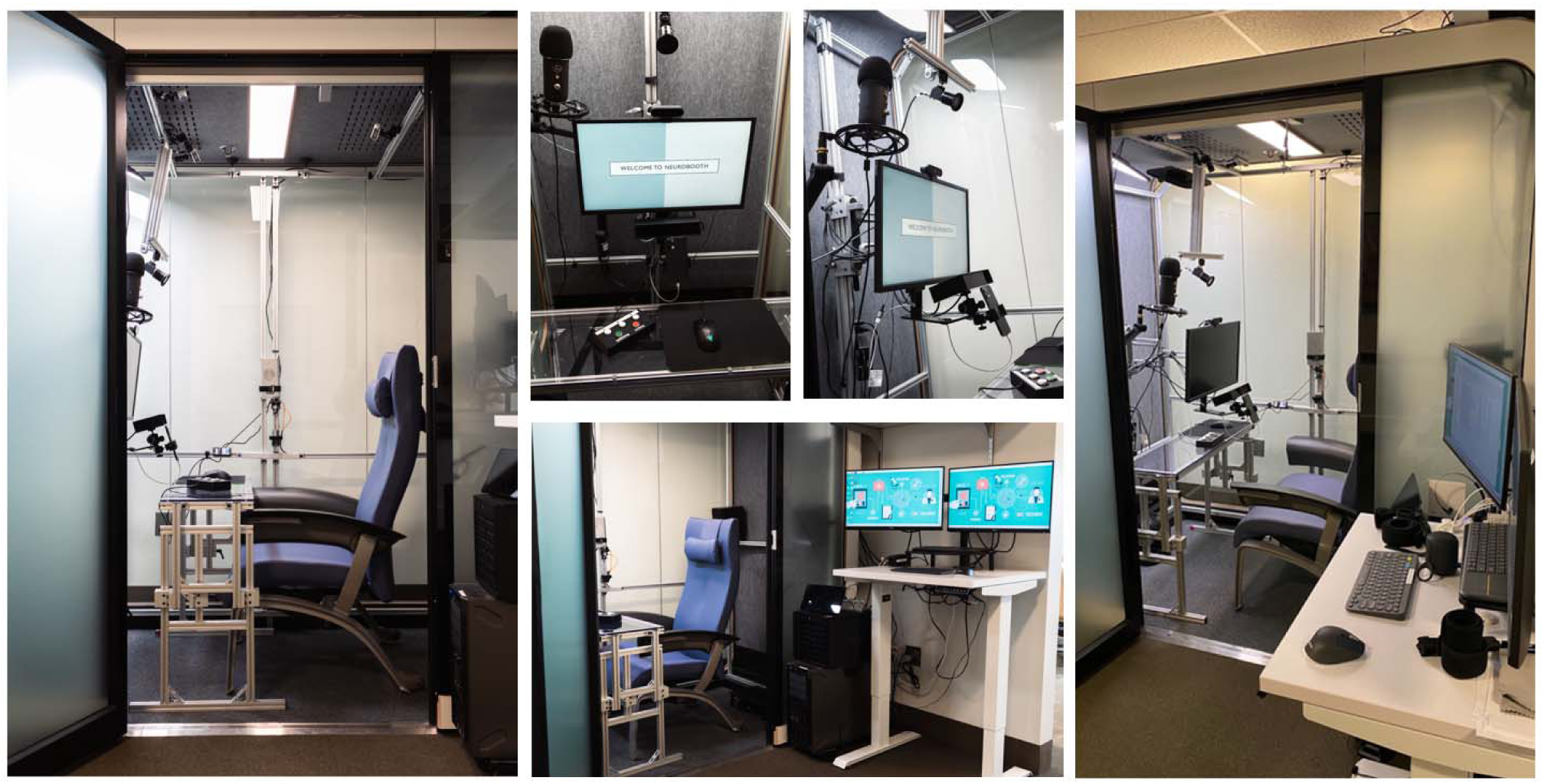
The Neurobooth, located in the MGH neurology clinic.

Between April 2022 and April 2024, data were collected during 782 sessions from 470 unique participants. Table 1 shows the clinical characteristics of these participants. A total of 213 of these participants have longitudinal data spanning up to 20 months (141 participated in two sessions, 45 participated in three sessions, and 27 participated in four sessions to date).

**Table 1.** Participant demographics and clinical diagnoses. Abbreviations: ARSACS—Autosomal recessive spastic ataxia of Charlevoix-Saguenay; CANVAS—Cerebellar Ataxia, Neuropathy, and Vestibular Areflexia Syndrome

| Participant diagnosis | N | Age in decades | Female % |
| --- | --- | --- | --- |
| Control | 82 | 2nd to 8th | 70 |
| Alzheimer's disease | 5 | 5th to 8th | 60 |
| Amyotrophic lateral sclerosis | 4 | 5th to 7th | 25 |
| ARSACS | 3 | 1st to 2nd | 100 |
| Atypical parkinsonism | 9 | 5th to 8th | 56 |
| CANVAS | 11 | 3rd to 7th | 45 |
| Corticobasal syndrome | 4 | 7th | 75 |
| Fragile X-associated tremor/ataxia syndrome | 4 | 5th to 7th | 50 |
| Friedreich's ataxia | 14 | 2nd to 7th | 79 |
| Frontotemporal dementia | 2 | 7th | 100 |
| Hereditary spastic paraplegias | 10 | 4th to 7th | 50 |
| Huntington's disease | 1 | 6th | 0 |
| Lewy body dementia | 3 | 6th to 8th | 100 |
| Mild cognitive impairment | 5 | 3rd to 8th | 80 |
| Multiple system atrophy - cerebellar type | 8 | 6th to 7th | 88 |
| Multiple system atrophy - parkinsonian type | 4 | 5th to 7th | 25 |
| Other neurological diagnosis | 36 | 3rd to 9th | 53 |
| Other ataxia | 61 | 2nd to 8th | 34 |
| Parkinson's disease | 132 | 4th to 8th | 33 |
| Primary progressive aphasia | 7 | 5th to 7th | 29 |
| Progressive supranuclear palsy | 14 | 6th to 8th | 79 |
| Spinocerebellar ataxia 1 | 2 | 3rd to 6th | 100 |
| Spinocerebellar ataxia 2 | 8 | 2nd to 7th | 63 |
| Spinocerebellar ataxia 3 | 24 | 2nd to 7th | 71 |
| Spinocerebellar ataxia 6<sup>†</sup> | 13 | 5th to 8th | 62 |
| Spinocerebellar ataxia 8 | 3 | 3rd to 5th | 67 |
| Spinocerebellar ataxia 27B | 1 | 5th | 0 |

<sup>†</sup>One subject with spinocerebellar ataxia 6 was concurrently diagnosed with spinocerebellar ataxia 8.

In addition to synchronized time-series data collected from a variety of sensors (Table 2), a corpus of contextual clinical data was collected to support development of clinically meaningful digital features. These data included both clinician-performed disease rating scales that were completed during the clinic visit and a battery of patient-completed surveys capturing information about daily function and quality of life.

**Table 2.**
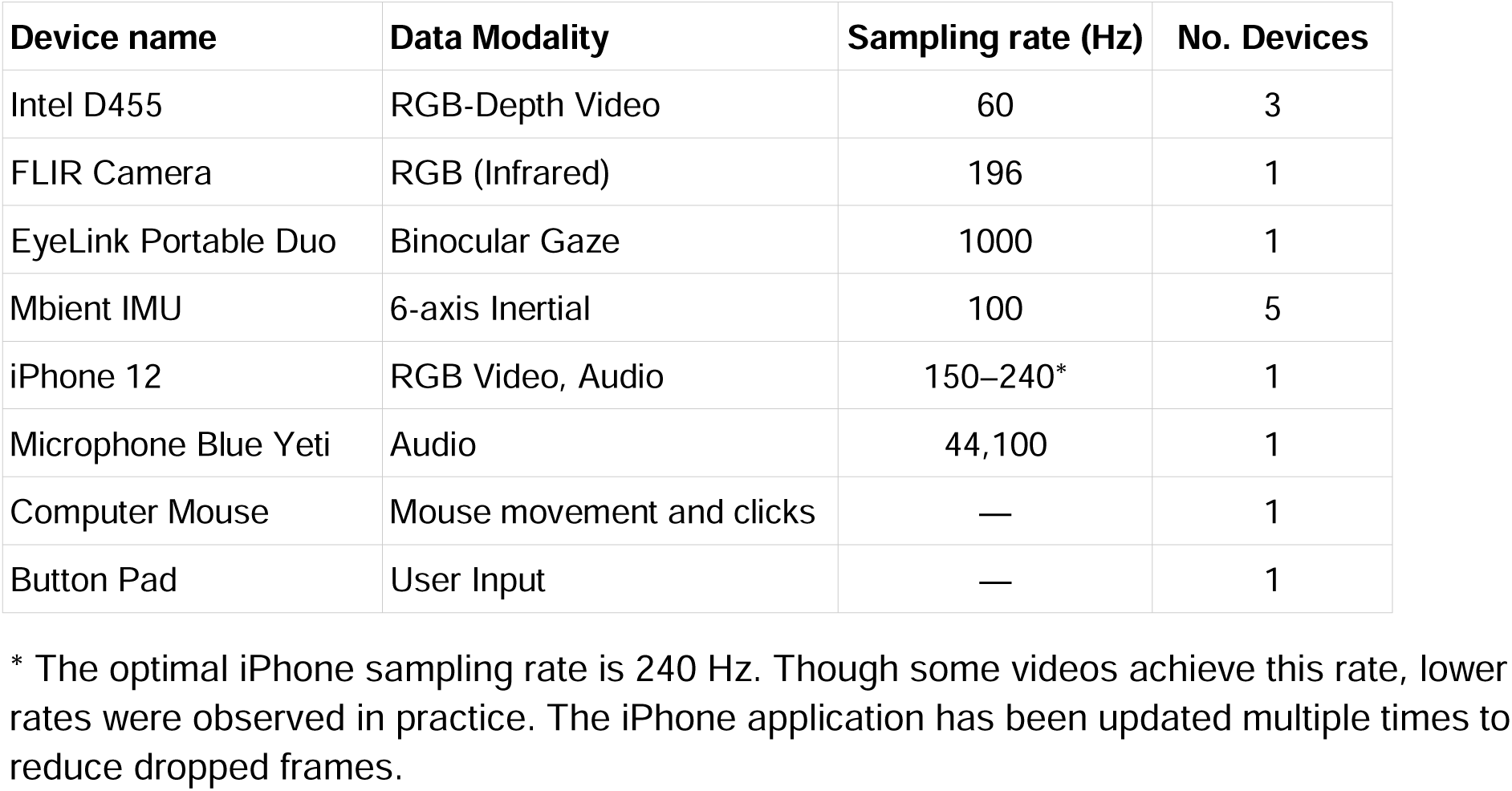
Data collection devices in the initial Neurobooth deployment. Data were captured from all devices during most tasks, with exception of computer mouse data, which were only captured during tasks that used the mouse as an input device. Data were not collected from the eye tracker or iPhone during the motor tasks because the arm holding the eye tracker and iPhone was repositioned during these tasks to create room for task performance.

| Device name | Data Modality | Sampling rate (Hz) | No. Devices |
| --- | --- | --- | --- |
| Intel D455 | RGB-Depth Video | 60 | 3 |
| FLIR Camera | RGB (Infrared) | 196 | 1 |
| EyeLink Portable Duo | Binocular Gaze | 1000 | 1 |
| Mbient IMU | 6-axis Inertial | 100 | 5 |
| iPhone 12 | RGB Video, Audio | 150–240* | 1 |
| Microphone Blue Yeti | Audio | 44,100 | 1 |
| Computer Mouse | Mouse movement and clicks | — | 1 |
| Button Pad | User Input | — | 1 |

\* The optimal iPhone sampling rate is 240 Hz. Though some videos achieve this rate, lower rates were observed in practice. The iPhone application has been updated multiple times to reduce dropped frames.

### Behavioral Tasks

Neurobooth data collection deployed a set of tasks spanning several neuromotor and neurocognitive behavioral domains—cognition, oculomotor function, fine and gross motor function, and speech—that are broadly relevant across neurodegenerative diseases and can be performed in a limited space and time setting (see Methods for task details). Several tasks involved coordination of multiple behavioral domains, taking advantage of the synchronous multimodal data capture supported by Neurobooth. The initial task set was designed such that it could be completed by most participants in 30-40 minutes without over-taxing patients on their clinic day, and to maximize the number of people who could participate on a given day. In the sections below, we provide a brief overview of the behavioral tasks and provide a data visualization for each task that highlights a potential disease-related behavioral pattern to motivate future population-level data analysis.

### Cognitive Tasks

Cognitive tasks included the digit symbol substitution test (DSST) and the multiple object tracking test (MOT). We used the validated structure from TestMyBrain.org^63^ to develop the cognitive tasks in Python.

The top row of Fig. 2 contrasts performance on a four-target, low-speed trial of MOT for a control versus a participant with corticobasal syndrome (CBS). The trajectory of gaze is shown in gray and the trajectory of the computer mouse is shown in blue, with cyan dots representing mouse clicks. The participant with CBS had less direct and longer gaze and mouse paths during the response period. One target was incorrectly selected, although their gaze and mouse trajectories indicate they considered the correct target. Though not shown in the figure, tracking gaze throughout the task could reveal different attention strategies for performing the task, which may be modified by neurological diseases^64,65^.

**Figure 2.**
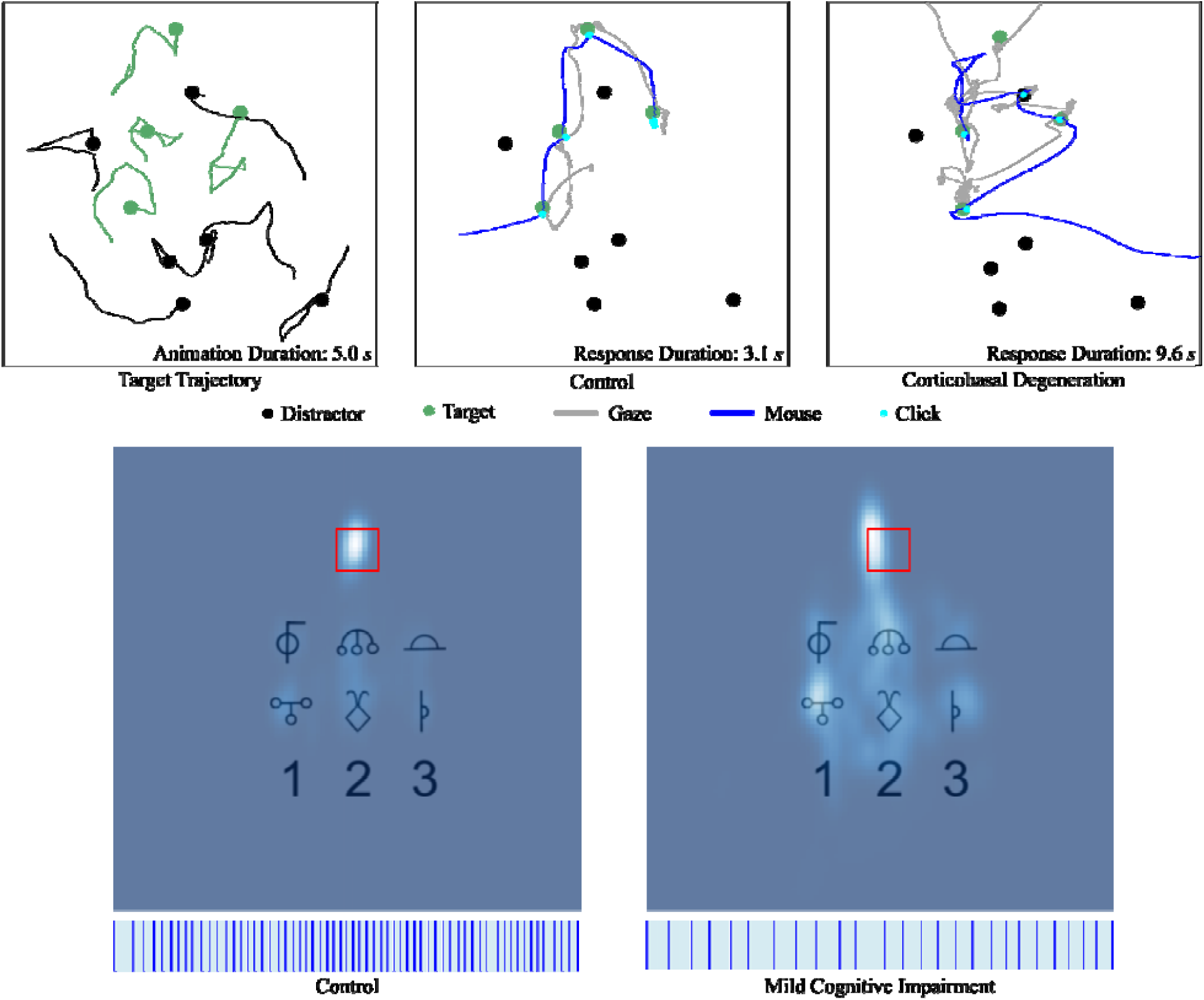
Illustrations of Multiple Object Tracking (MOT) and the Digital Symbol Substitution Test (DSST). (**top**) One trial of MOT with four targets and six distractors. The left panel displays the trajectory of each circle, with the dot indicating the final position. The middle and right panels display gaze and mouse trajectories from a control and participant with corticobasal syndrome, respectively. (**bottom**) Heat maps showing the relative gaze duration during DSST. The red square indicates the location of the target symbol. Lighter colors represent a longer time looking at a given region. The left panel shows a control and the right panel shows a participant with mild cognitive impairment. The vertical blue lines at the bottom represent the presentation of symbols during the one minute task duration, with shaded blue regions indicating the response time before the button press.

The bottom row of Fig. 2 compares a healthy control participant versus a participant with mild cognitive impairment (MCI) performing DSST. The heat maps show the density of gaze duration over the 60-second long task. The density in the left plot shows that the gaze of the control participant was largely fixated on the location of the stimulus symbol. In contrast, the cognitively impaired participant spent a significant time gazing at the key, as is apparent from the larger spread of the density in the heat map on the right. This shows that the attention of the participant with MCI was divided between the stimulus symbol and the key. The horizontal plots below the heat maps are a time series of events over the 60-second task. Vertical blue lines indicate when a new stimulus symbol appeared. Shaded blue regions indicate the length of the response times. The control participant registered 61 responses compared to 25 responses from the MCI participant. Differences in the thickness of the blue shaded regions provide a visual indication of the variance in response times. Both the control and the MCI participant were 100% accurate in these sessions.

### Oculomotor Tasks

Oculomotor tasks included tasks that include a visual target (horizontal smooth pursuit, gaze holding, horizontal saccades, and vertical saccades) and gaze fixation without a target. Fig. 3 compares the gaze during each task between healthy controls and participants with neurologic disorders. For smooth pursuit, the individual with multiple system atrophy had saccadic pursuit, with more pronounced saccadic pursuit when tracking the object moving rightward. For horizontal saccades, the person with Gordon-Holmes syndrome had hypermetric saccades in addition to gaze-evoked nystagmus during fixations. For vertical saccades, the individual with progressive supranuclear palsy had slowed and segmented saccades and evidence of ‘round the houses’ sign^66^ (curved trajectory apparent with concurrent movement in the horizontal direction). For gaze holding, the individual with cerebellar ataxia, neuropathy, and vestibular areflexia syndrome (CANVAS) exhibits downbeat nystagmus that varies in intensity depending on direction of gaze. For fixation without a target, the individual with spinocerebellar ataxia (SCA) type 3 (SCA-3) has vertical drift of gaze, with corrective saccades, and superimposed nystagmus.

**Figure 3.**
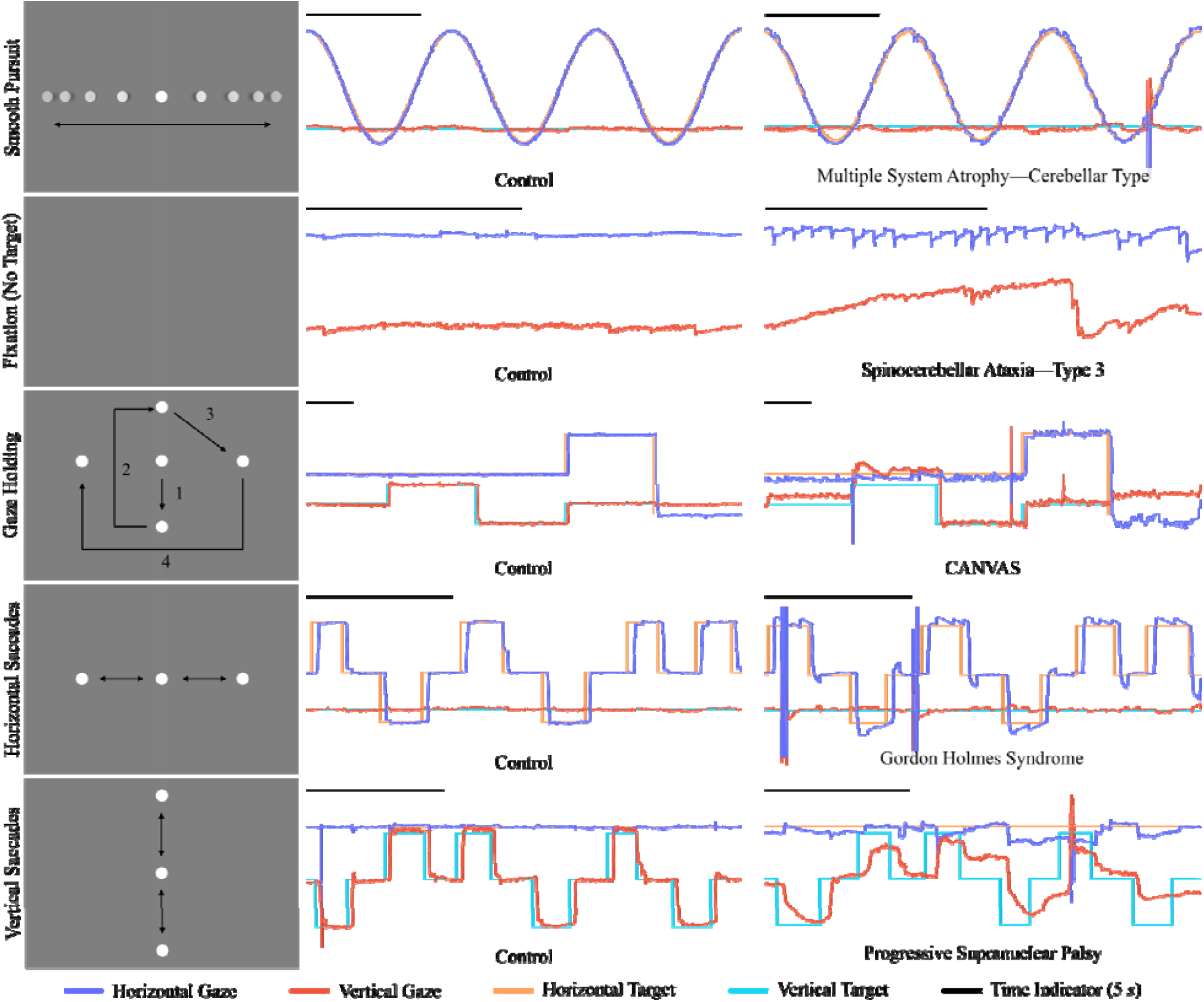
Illustrations of oculomotor tasks accompanied by example gaze time-series. Arrows and numbers indicate the motion of the visual target and are not shown to participants. Only a single visual target is displayed throughout each task. All control time-series were obtained from a single data collection session (i.e., from a single participant). Horizontal and vertical gaze positions are plotted on the y-axis with time represented on the x-axis. The y-axis values representing horizontal gaze increase with eye movement to the far right of the screen, whereas y-axis values representing vertical gaze increase with eye movement to the bottom of the screen.

### Motor Tasks

Motor tasks included finger-to-nose, foot tapping, rapid alternating hand movements, sit-to-stand, and the Hevelius mouse clicking task^58,59^. Fig 4 shows ten seconds of data collected during each task for a healthy control participant and an individual with a neurological disorder.

**Figure 4.**
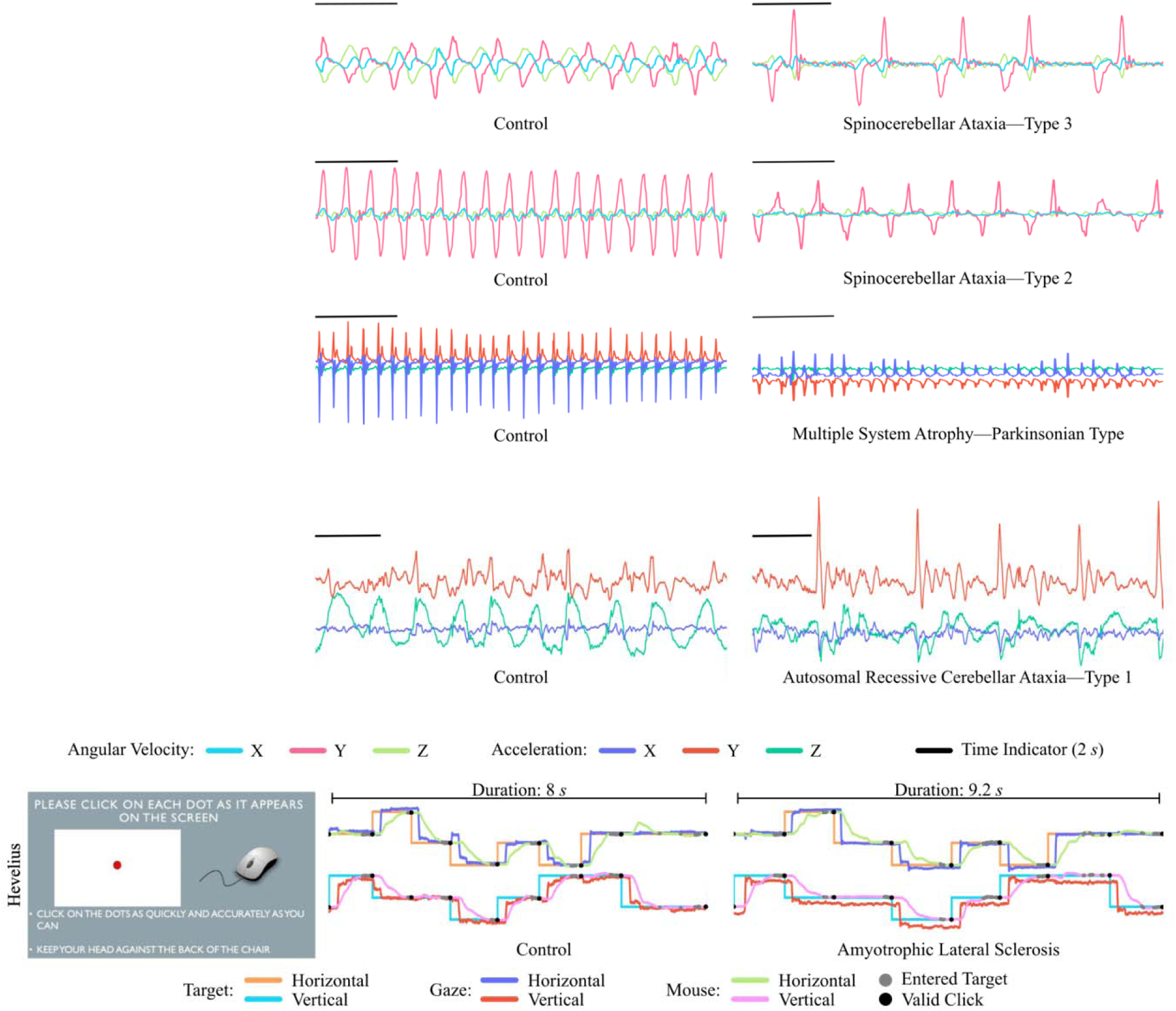
Illustrations of motor tasks. The finger-to-nose, alternating hand, foot tapping, and sit-to-stand tasks are accompanied by example triaxial inertial time-series. For the foot tapping task, the ankle sensor was oriented differently for the two displayed time-series. For the Hevelius computer mouse task, the example data are represented in screen coordinates. Horizontal screen coordinates increase from left to right. Vertical screen coordinates increase from top to bottom. The target moved immediately after a valid mouse click. It was possible for the mouse to enter the target multiple times prior to a valid click.

Finger-to-nose data from an individual with spinocerebellar ataxia type 3 (SCA-3) and rapid alternating hand movements data from an individual with SCA-2 demonstrate reduced speed and variability in amplitude and rhythm compared with the healthy control participant (Fig. 4).

Foot tapping data from an individual with multiple system atrophy, parkinsonian type (MSA-P) exhibits smaller movements and variability in amplitude compared with the healthy control participant (Fig. 4).

Sit-to-stand data (lumbar sensor) from an individual with autosomal recessive cerebellar ataxia type 1 (ARCA-1) shows less regular and lower intensity movements in the dorso-ventral axis and sharp peaks in the rostro-caudal axis compared to the smoother movement profile seen in the healthy control example.

On the computer mouse task, an individual with amyotrophic lateral sclerosis (ALS) took longer to perform the task and exhibited mouse movements that were less smooth and had more target re-entries. The presence of eye tracking data alongside mouse movements and stimulus onset provides an opportunity to assess how eye-hand coordination is affected by different motor disorders (Fig. 4).

### Speech Tasks

Speech tasks included sustained phonation, diadochokinetic tasks, and passage reading^67,68^. Fig. 5 compares recorded audio and gaze (for passage reading) for healthy control participants and individuals with neurologic disorders. For sustained phonation, in comparison with the control participant, a participant with Parkinson’s disease had decreasing loudness as the task progressed. For the diadochokinesis task, the individual with SCA-6 exhibited variability in amplitude and rhythm, as well as slowed consonant production. For passage reading, an individual with Friedreich’s ataxia had slowed speech and longer pauses during speech production, the presence of smaller saccades, and an increased frequency of regressions in gaze during reading.

**Figure 5.**
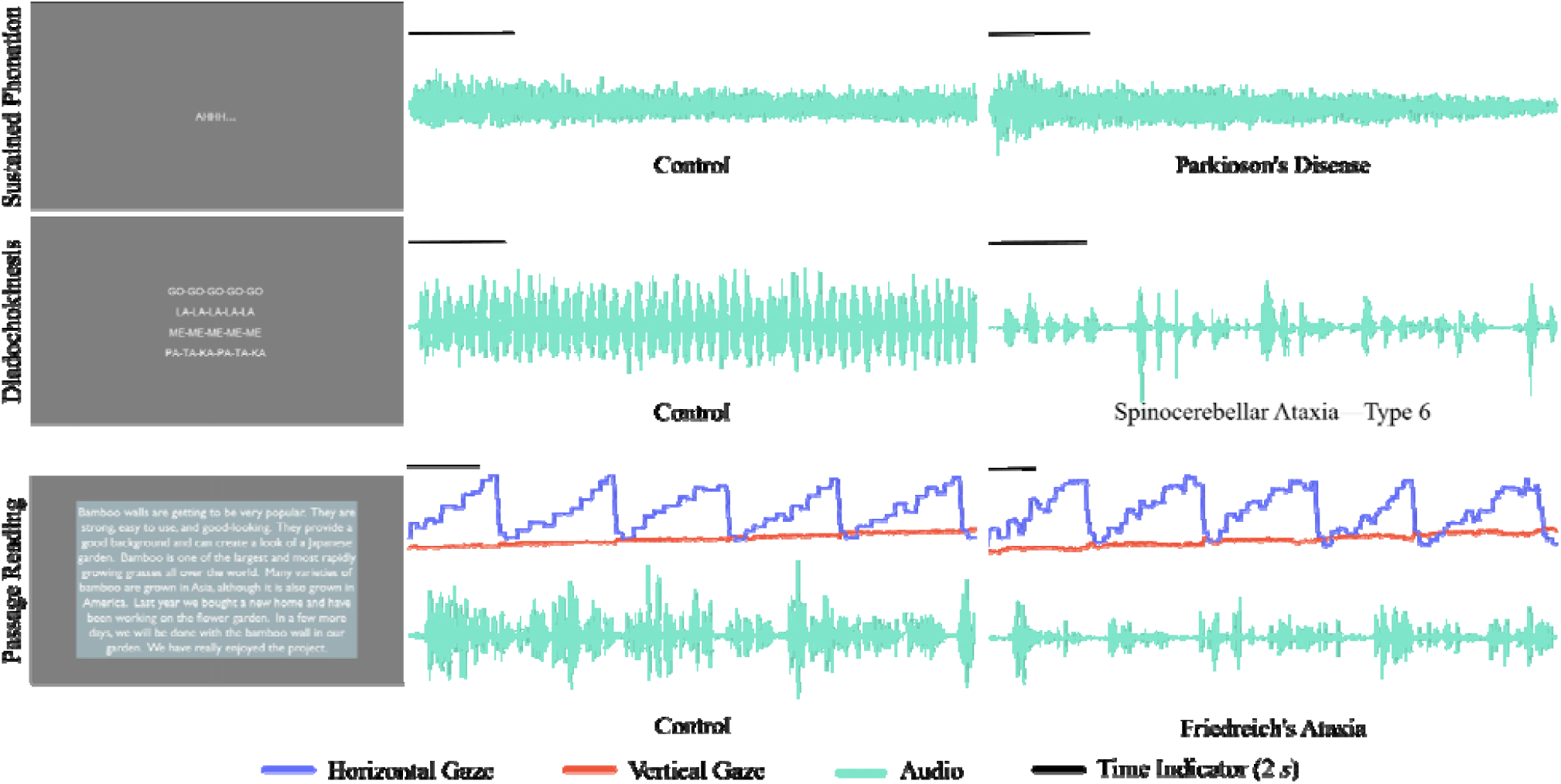
Illustrations of the speech tasks - sustained phonation (top), diadochokinesis tasks (middle), and passage reading (bottom). For diadochokinesis tasks, only a single line of centered text is displayed per task. The illustrated diadochokinesis data are from the “Go-Go-Go” task. For passage reading, the y-axis values representing horizontal gaze (blue curve) increase with eye movement to the far right of the screen, whereas y-axis values representing vertical gaze (red curve) increase with eye movement to the bottom of the screen.

### Study Feedback

To collect participants’ feedback on the Neurobooth system and their overall study experience, we administered a standardized 10-item System Usability Scale (SUS)^69^ and a Study Feedback survey created by our team after each visit. A total of 581 responses were captured from 363 unique participants.

Each item in the SUS questionnaire was rated from 1 to 5 on the Likert scale (1=strongly disagree and 5=strongly agree). To interpret the score, we calculated the sum of the score contributions for each item. For items 1, 3, 5, 7 and 9 (odd-numbered, positive statements), the score contribution was the scale position minus 1. For items 2, 4, 6, 8 and 10 (even-numbered, negative statements), the score contribution was 5 minus the scale position. Next, we multiplied the total by 2.5 to obtain the overall System Usability value, which ranges from 0 (very poor perceived usability) to 100 (excellent perceived usability)^69^. Mean SUS score from Neurobooth respondents was 78.4 with a standard deviation of 16.4. This meets the benchmark score of 68.05 for digital health applications as described in the current literature^70^.

In the Study Feedback survey, we assessed participants’ willingness to complete the full Neurobooth assessment again in the future. The majority of responses indicated yes, they would be interested (81.1%) while some expressed willingness to complete a shorter version of the assessment (11.8%). A smaller proportion of responses indicated participants were unsure (3.5%), they would probably not be willing to participate again (2.6%), or they would definitely not participate again (1.0%). We also surveyed participants on their most and least enjoyed study tasks. 42.0% of responses indicated cognitive tasks as their most enjoyed, followed by 21.7% for motor tasks. For least enjoyed tasks, 36.7% indicated none (they enjoyed all of the activities), followed by 26.8% for speech tasks. Lastly, participants were given an optional, free-text comment box to report any neurological symptoms of theirs they felt were not captured during testing. Participant responses, ordered from most to least frequent, included balance, gait, visual impairments (blurred vision, double vision, oscillopsia), neuropathy, weakness, handwriting, intermittent symptoms/general fluctuations throughout the day, dizziness, using stairs, reading comprehension, and lower extremity coordination during weight bearing activities.

## Discussion

In the first two years of the project, data from hundreds of individuals have been collected using Neurobooth, demonstrating the feasibility of collecting high throughput, multimodal digital data from diverse disease populations in the outpatient neurology clinic setting. Data visualization at the individual level indicates the ability to capture key neurological signs in the range of populations studied. This granular and comprehensive dataset provides an opportunity to detect and quantify neurological signs, characterize early disease features and change over time, and identify the presence of neurological subgroups and subphenotypes. Developing an open source platform for standardized data collection that can be used at other institutions enables the possibility of generating even larger datasets in aggregate. These datasets, combined with sophisticated computational modeling techniques, will lead to a more complete understanding of how neurological diseases affect behavior.

Neurobooth consists of a wide array of devices, including cameras, wearable sensors, an eye tracker, microphone, computer mouse, button board, and a mobile phone (with integrated camera and microphone). These devices, along with a broad set of behavioral tasks, enable a comprehensive capture of behavior relevant across neurological conditions. In addition to supporting single-domain analyses in a larger sample than prior studies, Neurobooth provides multimodal, time-synchronized data enabling new opportunities for the identification of less obvious patterns relating to the coordination of multiple motor domains. For example, multimodal modeling^71–73^ (e.g., eye movements and mouse movements during the computer mouse task; or speech, eye movements, and face movements during passage reading^74^) could enable discovery of new disease signs related to coordination of different behavioral domains that were not previously recognized or measurable by human assessors. Recent digital phenotyping efforts for screening of neurodevelopmental disorders highlights the importance of multimodal data^75,76^. The collection of contextual gold standard clinician rating scale data and patient reported measures of function enables validation of existing or newly discovered digital behavioral patterns or signs. Furthermore, the collection of longitudinal data, beginning in some individuals in the presymptomatic stage of disease, supports understanding how a digitally captured behavioral pattern begins and evolves over time. Although the diagnosis used for analysis will be the treating neurologist’s diagnosis, in cases where a definitive diagnosis can only be made based on neuropathology (e.g., progressive supranuclear palsy), attempts will be made to follow individuals until autopsy when possible.

Neurobooth incorporates a broad array of devices, and also allows for the integration of new device types. The system can be extended to include devices to measure strength or to measure physiological signals synchronously with behavior, such as respiration, electrocardiography (ECG), and electroencephalography (EEG). This flexibility makes it possible to broaden investigations while maintaining standardized data capture that allow for aggregation. Conversely, a system with a reduced set of devices can be deployed to reduce cost and allow data capture in a variety of settings. To this end, we are currently working to make it possible for a reduced version of Neurobooth to run on a single laptop and enable data collection in the home setting with consumer grade devices. In addition to device-flexibility, there is flexibility to change or remove existing behavioral tasks and to include additional tasks in a recording session. Thus the system can also be adapted to facilitate research beyond neurological populations. One goal of the platform is to maintain an open source library of behavioral tasks and device integrations that can be drawn upon by any research group or clinical team.

There were challenges encountered in working with the range of disease populations and severity levels, especially given the time-constrained setting in the neurology clinic. Although Neurobooth is accessible for individuals who are wheelchair bound and would prefer to remain in their wheelchair for the session, the quality of eye tracking is sometimes reduced due to lack of adequate head support with some wheelchairs. In individuals with nystagmus or gaze holding difficulties, calibration and validation of the eye tracker can be time-consuming and occasionally unsuccessful after multiple attempts. Modifications such as inserting a cushioned headrest and adjusting the height of the computer monitor have improved eye tracking quality. The Mbient wearable inertial sensors that stream data continuously over Bluetooth to support synchronization, could at times lose connectivity resulting in lost data. Changing the location of Bluetooth receivers and automating wearable device resets prior to motor tasks improved data capture. Individuals with cognitive impairments in some cases had difficulty understanding and following task instructions, requiring repetition of the instructions or skipped tasks, and lengthening the duration of the session. In particular, the Multiple Object Tracking (MOT) task could take substantially longer for individuals with cognitive deficits to complete. An interim software update was made to automatically terminate the task after the easier trials if the task was taking extended time. Future studies focused on a reduced range of neurological populations could benefit from modifying the task set to optimize for feasibility and phenotypic capture.

In summary, Neurobooth is a flexible research platform to collect multimodal quantitative behavioral data, with the goal to advance computational investigations of human behavior. The open source platform aims to reduce barriers for collecting standardized behavioral data and allow large aggregate datasets to emerge over time. These large multimodal datasets can then fuel discovery of new disease characteristics and result in a deeper understanding for how brain diseases give rise to changes in behavior. Ongoing work aims to increase the scale of the platform by enabling the system to run the same behavioral tasks using a subset of devices connected to a single laptop computer. An additional avenue of active research is the creation of data visualizations and reports that can summarize the complex multimodal digital data and are useful for patients and clinical providers.

## Methods

### Neurobooth Hardware

Given space constraints (and in order to maintain a quiet and private environment), MGH Neurobooth was built within a physical booth designed to be a small meeting space for 2–4 people (Framery Q model, Framery Oy., Finland). The frame has acoustic insulation and ventilation, antimicrobial surfaces, and is wheelchair-accessible. An 80/20 aluminum rail structure was installed inside the booth to mount an adjustable computer monitor and a broad array of sensors. A sprinkler was installed in the booth ceiling to comply with hospital safety regulations and a privacy film was applied to the glass walls facing the clinic. Additional light panels were installed inside the booth to increase the luminance of recorded videos and an intercom system was installed to facilitate communication between the participant inside the booth and research staff outside of the booth. A chair with a reclining back and adjustable headrest was added to improve participant comfort and eye tracking data quality. However, the chair can be removed for those who prefer to complete their session in their wheelchair. Fig. 1 illustrates the complete physical layout of the Neurobooth.

The devices included in MGH Neurobooth support data capture across several behavioral domains (Table 2). Participants interacted with a button board to advance through task instructions and to provide responses during cognitive tasks. The computer mouse was used to capture arm movement and button clicks for a subset of tasks (i.e., MOT, Hevelius). Participant motion was captured using five wearable Mbient inertial sensors placed on the wrists, ankles, and lumbar region streaming triaxial acceleration and angular velocity. Three Intel RealSense D455 RGB-Depth cameras were located on the ceiling of the booth forming a triangle to enable estimation of body landmark positions in 3D space. High-quality eye tracking data were collected using an EyeLink Portable Duo placed below the monitor. An industrial-grade, high-speed FLIR camera was positioned above the monitor to support capture of fast facial movements. The back-facing camera of an iPhone model 12 running a custom data capture application was positioned below the eye-tracker to support capture of facial movements and eye movements^40,41^. Both a high-quality Blue Yeti microphone and the iPhone captured audio (i.e., participant speech).

### Neurobooth Software

A custom Python-based library, which we call the Neurobooth Operating System (NeuroboothOS), coordinates behavioral stimulus delivery and data collection. NeuroboothOS provides a graphical user interface (GUI) for the operator to initiate and terminate data collection. Delivery of task instructions and stimuli to the participant is accomplished using PsychoPy^77^, an open source Python toolbox. This design enables researchers to create and deploy behavioral tasks with relative ease. Tasks and their associated devices are organized in collections and studies.Task variables along with device and sensor parameters are defined in a set of YAML configuration files so they can be readily adjusted. A script is provided to verify that the set of files defining a study is complete and consistent. Data collected from each device is synchronized using the lab streaming layer (LSL) library^78^. Logs for each session, task, and data file are generated in a PostgreSQL database. The same database contains participants’ clinical, demographic, and patient-reported information automatically ingested from REDCap^79^, an electronic data capture (EDC) system. An interactive web-based tool called the Neurobooth Explorer allows time series data to be visualized in real time after the session and can export visualizations. The time-series data figures in this manuscript were exported from the Neurobooth Explorer. All software is available on the neurobooth GitHub repository, https://github.com/neurobooth/.

### Contextual Clinical Outcomes

Clinical rating scales were performed by clinical providers as part of the routine clinical appointment. For participants with ALS, the ALS Functional Rating Scale-Revised (ALSFRS-R)^80^ was performed. For participants with Alzheimer’s disease, mild cognitive impairment and other memory disorders, the Montreal Cognitive Assessment (MOCA) was performed^81^. For participants with a form of parkinsonism, the MDS-Unified Parkinson’s Disease Rating Scale (MDS-UPDRS)^82^ was performed. For participants with ataxia, the Brief Ataxia Rating Scale (BARS)^83^, the Scale for Assessment and Rating of Ataxia (SARA)^84^, and components of the Modified International Cooperative Ataxia Rating Scale (MICARS)^83^ (arm, speech, and oculomotor sections) were performed.

We reviewed 42 patient-reported outcome measures (PROMS) used in neurodegenerative populations and selected a subset of 19 PROMS capturing information across the following domains: motor, speech, oculomotor, cognition, mood, participation in social activities, fatigue, sleep, quality of life (QoL), and environmental factors (Table 3). For the Neuro-QoL PROMS^85^, the short forms were chosen wherever possible to help minimize survey burden. PROMS available in the public domain were prioritized. In addition to existing PROMs, our team also created a survey assessing the frequency of falls over the past 3, 6, and 12 months. Participants with Parkinson’s Disease and ataxias completed disease-specific outcome measures, such as PROM-Ataxia^86^ and the Dysarthria Impact Scale^87^.

**Table 3.**
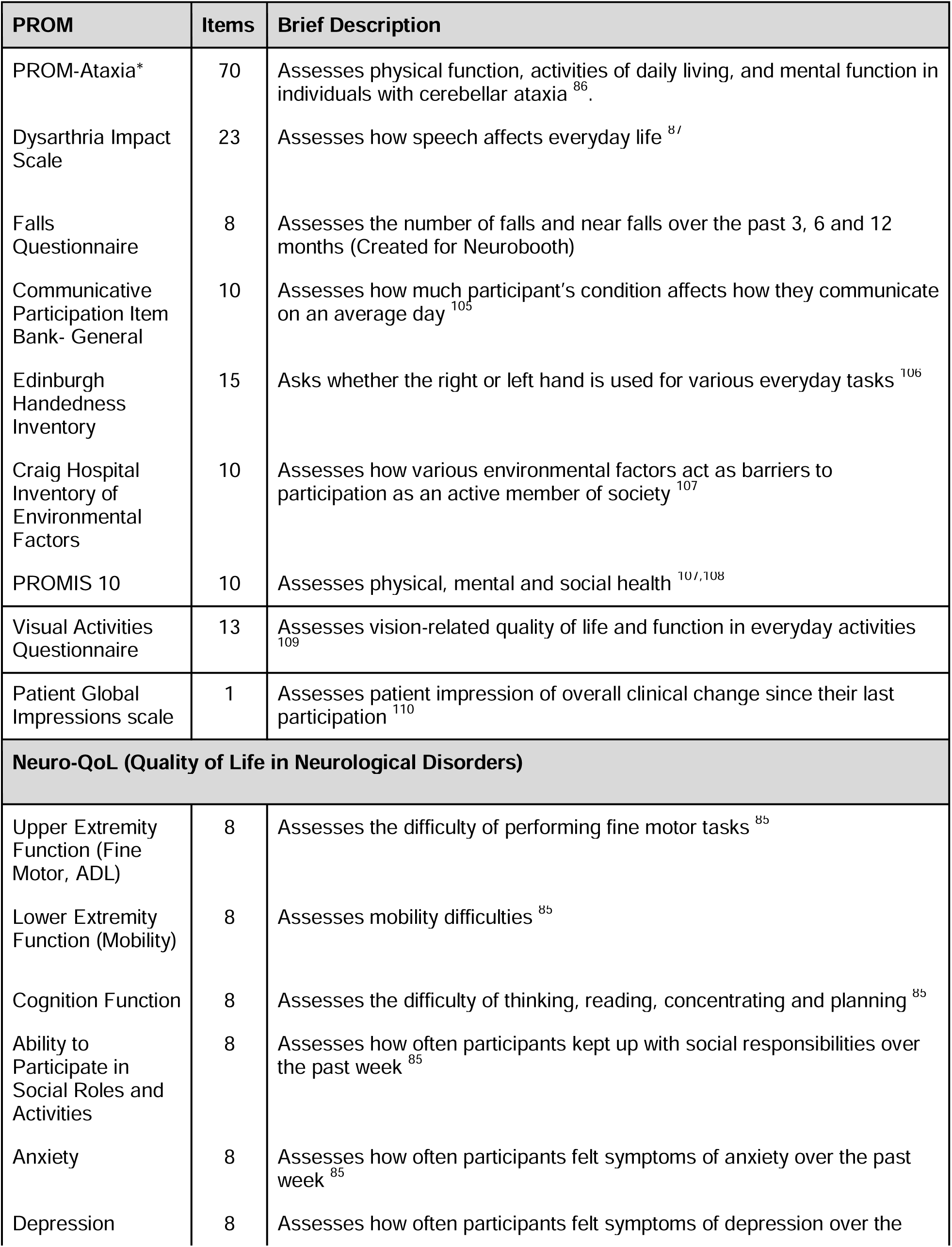
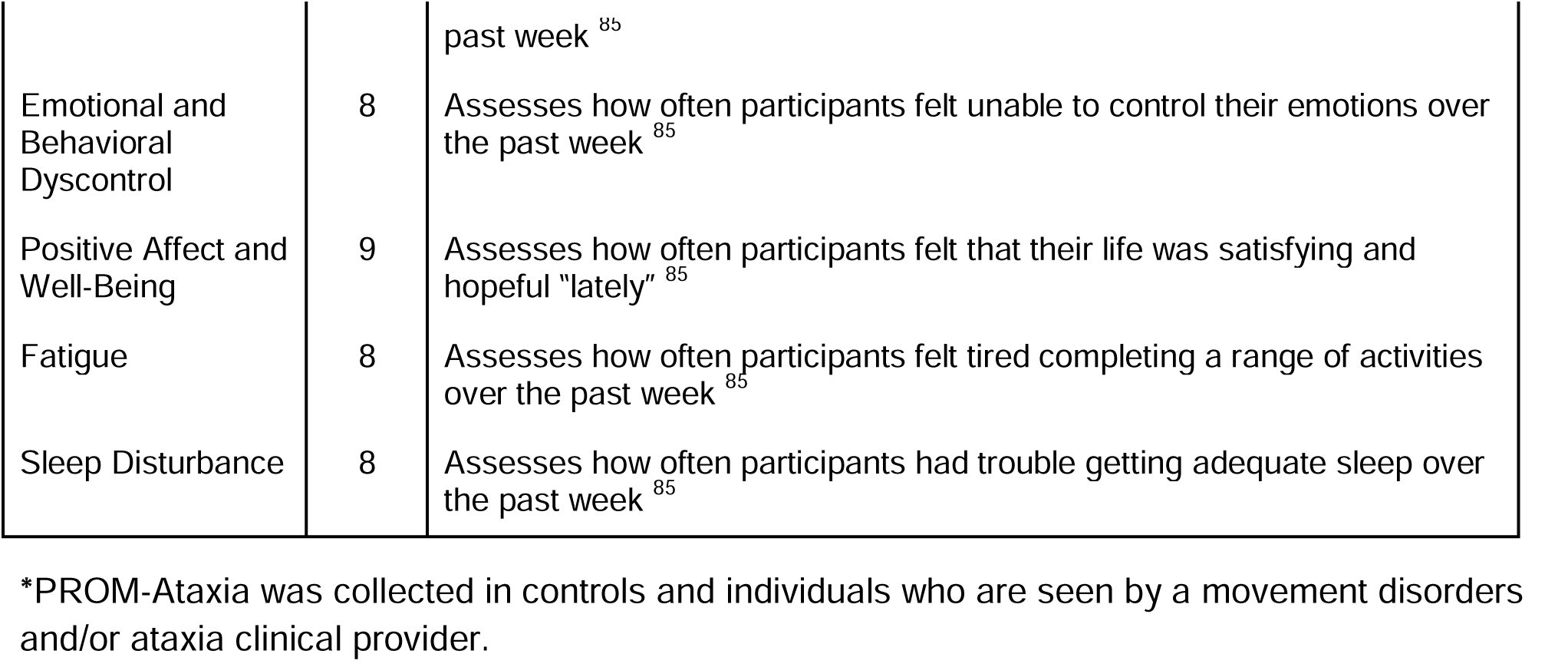
Descriptions of each collected patient reported outcome measure (PROM)

### Behavioral Tasks

#### Cognitive Tasks

The cognitive tasks were built in Python based upon the validated structure from TestMyBrain.org^63^. For the Digit Symbol Substitution Test (DSST)^88^, participants were shown six symbols arranged into three columns (Fig. 2). The columns were labeled “1”, “2”, and “3” such that each number was associated with two symbols. For each trial of DSST, a target symbol was shown to participants. Participants then attempted to choose the corresponding number using the button board. A new trial was immediately presented to participants once a button was pressed, with no indication of whether the selection was correct. Participants were given one minute to complete as many trials as possible. DSST assesses a large variety of cognitive domains including executive and visuoperceptual function, motor speed, attention, and associative learning.

For Multiple Object Tracking (MOT)^89^, participants were shown ten black dots (Fig. 2). Three to five of these dots were targets and the rest were distractors. The color of the target dots alternated between green and black for three seconds before reverting to black. All dots then simultaneously moved for five seconds. Once the dots were stationary, participants attempted to click the targets using the computer mouse in any order. Correct clicks were displayed as green and incorrect clicks were displayed as red. If the participant did not complete their clicks within one minute, then the trial was restarted. The full MOT task consisted of three blocks of six trials each (for a total of 18 trials). Each block corresponded to the number of targets (starting with three and ending with five) and successive trials within a block linearly increased the movement speed of the dots during the animation phase. (Dots moved at approximately 40 *px/s* in the first trial and 240 *px/s* in the last trial of each chunk.) The MOT task was significantly modified in May 2024, after the two-year period detailed in this manuscript. These modifications 1) ensured a more consistent stimulus delivery across sessions, 2) improved accessibility (i.e., larger fonts and color-blind-friendly colors), and 3) implemented early-stopping criteria for study participants who had notable difficulty performing the task based on long completion times in early trials. MOT assesses visual attention^89,90^.

#### Oculomotor Tasks

Prior to the oculomotor tasks, a five-point calibration task was performed to align the participant’s gaze with the screen coordinate system. At the beginning of each oculomotor task, participants were asked to maintain their head position against the back of the chair and to keep their head still and only move their eyes.

In the horizontal smooth pursuit task, participants tracked a small circular target as it moved horizontally across the screen for 30 seconds. The circle’s velocity varied in a sinusoidal manner, with zero velocity near the screen edges and maximal velocity (33.3 degrees/s) at the screen center. The task was included to assess the gain and smoothness of eye movements while tracking a slowly moving target stimulus, which can be impaired in neurological conditions^91–94^.

For the horizontal saccades task, participants tracked a circular target as it instantly appeared at the left, center, and right of the screen. The vertical saccades task was similar, with the target appearing at the bottom, center, and top of the screen. The circle was stationary between transitions for a short, randomized duration (uniformly distributed between 1–1.5 s). The horizontal and vertical saccades tasks assess multiple brain areas involved in the initiation and control of ballistic eye movements^95^.

In the fixation without a target task, participants stared at the center of a uniformly gray screen for 10 seconds. In the gaze-holding task, participants fixated on a target that moved between five different screen positions (i.e., center, down, up, right, then left). The circle was stationary in each position for 10 seconds before jumping to the next position. The fixation and gaze holding tasks were included to capture oculomotor signs such as nystagmus, saccadic intrusions, and drift^96^.

#### Motor Tasks

For the finger-to-nose task, participants were asked to extend their arm to about 90% of their reach to touch a circular target on the monitor and then return to their nose. This action was repeated for 15 seconds for each arm. The finger-to-nose task assesses upper extremity coordination, dysmetria, speed of movement, and tremor^97^.

In the repetitive foot tapping task, participants were instructed to repetitively tap their toes on the ground while maintaining heel contact with the ground. This action was performed for 10 seconds for each foot. For the rapid alternating hand movement task, participants were asked to raise their arm straight out in front of them and repetitively pronate and supinate their arm. This action was repeated for 15 seconds for each arm. These two tasks assess limb coordination and presence of bradykinesia^98^.

For the sit-to-stand task, participants were asked to stand up fully from a seated position and then sit back down as quickly as possible with their arms crossed across their chest. Participants were able to use the chair’s armrests if needed. The action was repeated 5 times. The sit-to-stand task assesses balance and lower extremity strength^99^.

In the Hevelius mouse clicking task^58^, participants were shown a single red dot on the screen and asked to click it using the mouse as quickly and as accurately as possible. After being clicked, the dot instantly moved to a new position on the screen. The task was divided into eight blocks, with each block presenting a sequence of nine dots. The size of the dot and the distance it moved on the screen remained the same within a block, but varied across blocks. It was previously observed that mouse trajectory, speed, and click characteristics were informative in quantifying ataxia and parkinsonism^58–60^.

#### Speech Tasks

For sustained phonation, participants were asked to say “ahh’’ for 10 seconds. Since participants were instructed to continuously produce sound, this task was intended to measure variations in frequency and amplitude, breath patterns, and vocal fluctuations during initiation and termination of the target vowel^100^. For the diadochokinesis task, participants were asked to repeat “go go go”, “la la la”, “me me me”, and “Pa-Ta-Ka” syllables as quickly and as many times as possible in 10 seconds. Prior to the sustained phonation and diadochokinesis tasks, participants were asked to take a deep breath and to continue the task until told to stop. These tasks are designed to capture speaking rate and vocal irregularities that may be present in many neurologic conditions^101^.

For the passage reading task, participants read aloud the Bamboo passage^102,103^, which is designed to include voiced consonants and phrase boundaries. Participants were asked to speak at their normal conversational volume and pace when reading the passage. Audio collected during the task can be used to assess speaking rate, phrase duration, and pauses^104^. Eye tracking allows assessment of eye movements during a naturalistic task and for assessment of eye-voice coordination^74^.

## Data Availability

All data produced in the present study are available upon reasonable request to the authors

## Data Availability

Data can be requested by qualified researchers by visiting https://neurobooth.mgh.harvard.edu/.

## Software Availability

Neurobooth software can be downloaded from our Github page: https://github.com/neurobooth/.

